# Assessing the key factors of quality of life for institutionalized elderly in Bangladesh

**DOI:** 10.1101/2025.06.13.25329612

**Authors:** Pradip Kumar Saha, Mohima Sharmin, Tasrima Trisha Ratna, Somaiya Islam, Masum Mridha, G M Mainuddin Chisty, Umme Salma, Kashfia Rahman, Shahriar Hasan

**Author notes:** **Corresponding author:** (PKS). (MM), (MS), (SH), (TTR), (SI), (GMC), (KR), (US).

## Abstract

**Background:** This study investigates the determinants of quality of life (QoL) among older adults residing in institutional settings in Bangladesh.

**Methods:** Utilizing a cross-sectional design, data were collected from 570 elderly participants across four institutional care facilities. Sociodemographic, physical health, dietary habits and comorbidity determinants were analyzed to determine their effect on QoL, measured through the OPQOL-35 scale.

**Results:** The findings indicated a strong association between lower QoL and age above 75 years, illiteracy, lower income, and absence of elderly allowances. Physical health and dietary habits like sleep, water, fruit intake with chronic diseases including skin disease, visual impairment, and arthritis were found to contribute heavily to poor QoL. Multivariate logistic regression indicated that the elderly above the age of 75 years were 5.14 times more likely to report poor QoL, and individuals with visual disorder were 2.72 times more likely to experience poor well-being. These findings identify the multifactorial determinants of the socioeconomic, health, dietary and comorbidity variables influencing institutionalized elderly QoL, indicating the necessity for specific interventions to improve QoL among the elderly population.

**Conclusion:** The research indicates that enhancing financial assistance, access to healthcare, and reducing gender inequality could positively contribute to the QoL of the elderly populations living at old homes in Bangladesh and similar low-resource countries.

## Introduction

The global elderly population is experiencing fast growth, which is indicating that the number of elderly aged 60 and older will reach 2.1 billion by 2050 [1]. This demographic shift is primarily driven by increasing life expectancy and declining birth rates, both of which will have substantial implications for healthcare systems worldwide [1]. It is estimated that by 2050, the elderly population will more than double, making up approximately 22% of the global population, thus imposing significant demands on healthcare and economic systems [2]. In particular, the elderly population (60 and older) is expected to increase to 1.5 billion by 2050 from the current population of 727 million [3], with the population of individuals aged 65 and older to account for a large segment of the population [3]. Consequently, public health systems will be put under more pressure to respond to the increase in the incidence of chronic diseases and the intensity of healthcare demands of the elderly [4].

Physical health, though the predominant influencing factor, is complemented by the emotional, mental, and social factors that contribute to QoL. Chronic diseases, mobility problems, or decline in cognitive functions can all impact physical health and lower QoL directly [5]. At the same time, mental illnesses like depression, anxiety, and loneliness profoundly affect the life satisfaction of elderly individuals [6]. Social relationships and participation in community activities further contribute to improved mental well-being and QoL [7]. Physical and mental well-being is further affected by the environment, such as the safety of the area where one stays, highlighting the necessity of an integrated approach to enhancing QoL among the elderly [6].

Furthermore, institutional care can provide the highly structured environments that support monitoring of the health status of individuals and social interaction, the availability of resources in the low- and middle-income countries (LMICs) tends to prevent the effectiveness of such care [8–10]. Market-led models of care of the elderly, for instance, lead to inequities in China, where high-quality services are unaffordable to low-income elderly individuals [11].

In Bangladesh, taking care of the elderly within the family has been a key part of the culture. However, rapid socioeconomic changes, demographic transitions, poverty, and evolving social norms have eroded this system, leading to a shift toward institutional care for older adults [12,13]. In spite of the resistance to institutionalization, the growing number of old age homes indicates an accommodation response to the problems that aging poses, where millions of elderly face lack of financial support, poor access to healthcare, and social insecurity [13]. New possibilities and challenges arise with the development of institutional care amidst the changing social scenario of Bangladesh [14].

As Bangladesh confronts this change, models of care that balance tradition with the realities of the day are needed urgently. This calls for large scale research using higher sample sizes with more sophisticated methodologies to feed into policy and care practice decisions. It is imperative to comprehend the determinants of QoL among the elderly who live in institutional settings, old homes, in particular, so that improved well-being can be promoted for this group of individuals. Sociodemographics, including age, education, and financial status, along with physical health status, as well as comorbidities, strongly impact QoL among this group of individuals. Environmental factors, particularly those of an institutional nature are critical to the elderly person’s overall well-being and health as well. This research aims to supplement current knowledge gaps through the provision of an exhaustive review of the determinants of QoL among elderly residents of old homes based in Bangladesh, the ultimate purpose of which is to inform policy and improve the approach to care.

## Methods

### Study Population

Cross-sectional design was used in this study to explore the determinants of the quality of life (QoL) of older adults at old homes in Bangladesh. The study was conducted from December 2023 to November 2024. Data collection was done among elderly participants who lived at the four conveniently selected sampling sites, which included the Old Rehabilitation Center at Gazipur, the Bangladesh Association for the Aged and Institute of Geriatric Medicine at Agargaon, Apon Vubon at Uttara, and Subarta Trust at Manikganj. Recruitment was carried out based on the criteria of including individuals who are 60 years or older, regardless of gender, to ensure a good coverage of the elderly population. This allowed for a comprehensive analysis of the questions under research.

### Ethical consideration

This research followed the ethical guidelines of the Declaration of Helsinki. This research also received an approval from the IRB of the Bangladesh Health Professions Institute (IRB/10/2023/781). Informed consent was obtained from all participants before they participated in this study. The participants were fully aware of the study’s objectives, methodology, and procedures. Participants had the full right to withdraw the interview at any time. Confidentiality and anonymity were strictly maintained. For this study no physical specimens were collected. Data were gathered solely through the standardized questionnaire.

### Sampling Process

Total 570 elderly people participated in the study. Participants were selected using a simple random sampling method. Those who provided written informed consent were invited to participate. If a participant declined, the next eligible person in the sequence was approached. This process ensured an unbiased selection and minimized non-response bias, contributing to the study’s overall validity.

### Survey Instrument

A structured questionnaire was used to collect the data. This questionnaire consisted four key sections. Each section was designed to capture different aspects of the participants’ sociodemographic, physical health, dietary habits & comorbidity status.

### Independent variables

Independent variables included participants’ age (60 to 75 years and above 75 years), marital status (married and unmarried), education status (illiterate, primary, secondary and above), monthly family income (below 25,000 BDT, 25,000-49,999 BDT, 50,000 and above BDT), monthly allowance (no and yes), body mass index (underweight, normal, overweight, and obese), water consumption (<8 glasses and 8 or more glasses), inclusion of vegetable in diet (always/sometimes and never), inclusion of fruits in diet (always/sometimes and never), inclusion of fish/meat/egg/lentil in diet (always/sometimes and never), inclusion of milk/milk products in diet (always/sometimes and never), sleep duration (< 8 hours and 8 or more hours), diabetes (present and absent), cardiovascular disease (present and absent), hypertension (HTN) (present and absent), respiratory disease (present and absent), thyroid disease (present and absent), urological disease (present and absent), dental disease (present and absent), arthritis (present and absent), visual disorder (present and absent), skin disease (present and absent), and low back pain (present and absent).

### Outcome variable

The OPQOL-35 questionnaire was used to assess the overall QoL of participants. This tool, developed by Ann Bowling [15]. This questionnaire employed a 5-point Likert scale and it ranged from "strongly agree" to "strongly disagree". The total score ranges from 35 to 175, with higher scores indicating better QoL. The score was categorized as follows: less than 99 = "bad as can be," 100-119 = "bad," 120-139 = "optimum," 140-159 = "good," and 160-175 = "good as can be". For the purpose of analysis, the original 5 categories of score were combined into two broader categories. The "Bad" and "Bad as can be" categories were converted into poor QOL. The "Optimum", "Good", and "Good as can be" categories were converted into Good QOL. To ensure validity, the questionnaire was forward and backward translated, and pilot test was conducted with 30 elderly individuals from the same demographic. The reliability of the questionnaire was verified by a Cronbach’s alpha of 0.81, which indicated a strong internal consistency.

### Data Analysis

For this study, STATA version 19 was used to analyze the data. Descriptive statistics were first employed to assess the data distribution. To examine the relationships between socio-demographic variables and QoL, Pearson’s chi-square test or Fisher’s exact test was used. To identify the risk factors which were associated with poor QoL, a binary logistic regression analysis was done. A bivariate logistic regression was conducted initially, followed by multivariate logistic regression using a purposeful selection method to control for confounding variables. The logistic regression model was specified as:

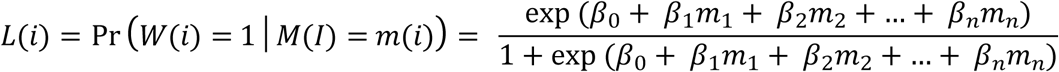

Where, *W(i)* = is the binary dependent variable for the *i*th participant:

*W(i)* = 1 if the participant reports a poor quality of life (QoL = “Bad” or “Bad as can be”)

*W(i)* = 0 if the participant reports a good quality of life (QoL = “Optimum”, “Good”, or “Good as can be”).

*β_0_* is the intercept of the model. *β_1_*, *β_2_*,…, *βn* are the coefficients for each covariate, indicating the association of each independent variable with the likelihood of reporting poor QoL. *m_1_,m_2_,…,m_n_* are the independent variables (covariates), such as: Age (e.g., 60–75 years vs. above 75 years), Gender (Male vs. Female), Education (Secondary and above vs. Illiterate, Primary), Monthly Family Income (e.g., 50,000 BDT and above vs. Below 25,000 BDT, 25,000 to 49,999 BDT) etc.

Hosmer and Lemeshow goodness-of-fit test was performed to evaluate the model’s fit, which demonstrated a chi-square value of 14.25 with 8 degrees of freedom (P = 0.075), indicating that the model fit the data well. to define statistical significance a p-value of < 0.05 was used in all analysis.

## Result

This study found that the average age of the participants was 64.86 years, with males exhibiting a slightly higher average age compared to females. Table 2 demonstrates, the majority of participants fell within the age range of 60-75 years (95.6%), with a greater proportion of females within this group (98.5%). Regarding marital status, the findings suggest that males were more likely to be married (91.5%) than females (57.6%). Hence, the data also revealed that females were more likely to be illiterate (60.5%) and possess lower levels of education compared to males (29.3%). In terms of income, the study found that a significant proportion of participants earned less than 25,000 BDT per month (43.2%), with no statistically significant difference between males and females. Nevertheless, the findings showed that females were more likely not to be receiving any monthly allowance (79.5%) than males. Physical health was found to be such that the greatest number contributed to the category of having a normal body mass index (BMI) (59.6%). Females showed more likelihood of obesity (10.7%) than males (4.1%). In addition, the research indicated that most of the subjects consumed more than 8 glasses of water daily (54.9%), with no distinguishing difference between the sexes. On the issue of nutrition, the results indicate that almost all the subjects consumed vegetables (98.9%), though the females ate milk-containing foods less frequently (76.1%) than the males (81.1%). The study also examined the prevalence of various health conditions among the participants. The results indicate that males were more likely to experience cardiovascular disease (38.4%), urological disease (35.3%), and respiratory disease (35.1%) compared to females. Conversely, females were more likely to experience arthritis (25.9%) compared to males. In addition, the study found the association of gender and aging (*P* = 0.011), marital status (*P* <0.001), Education (*P* <0.001), monthly allowance (*P<0.025*), BMI (*P* = 0.003), inclusion of fish/meat/egg/lentil in diet (*P* = 0.007), dental disease (*P* = 0.019).

**Table 1.**
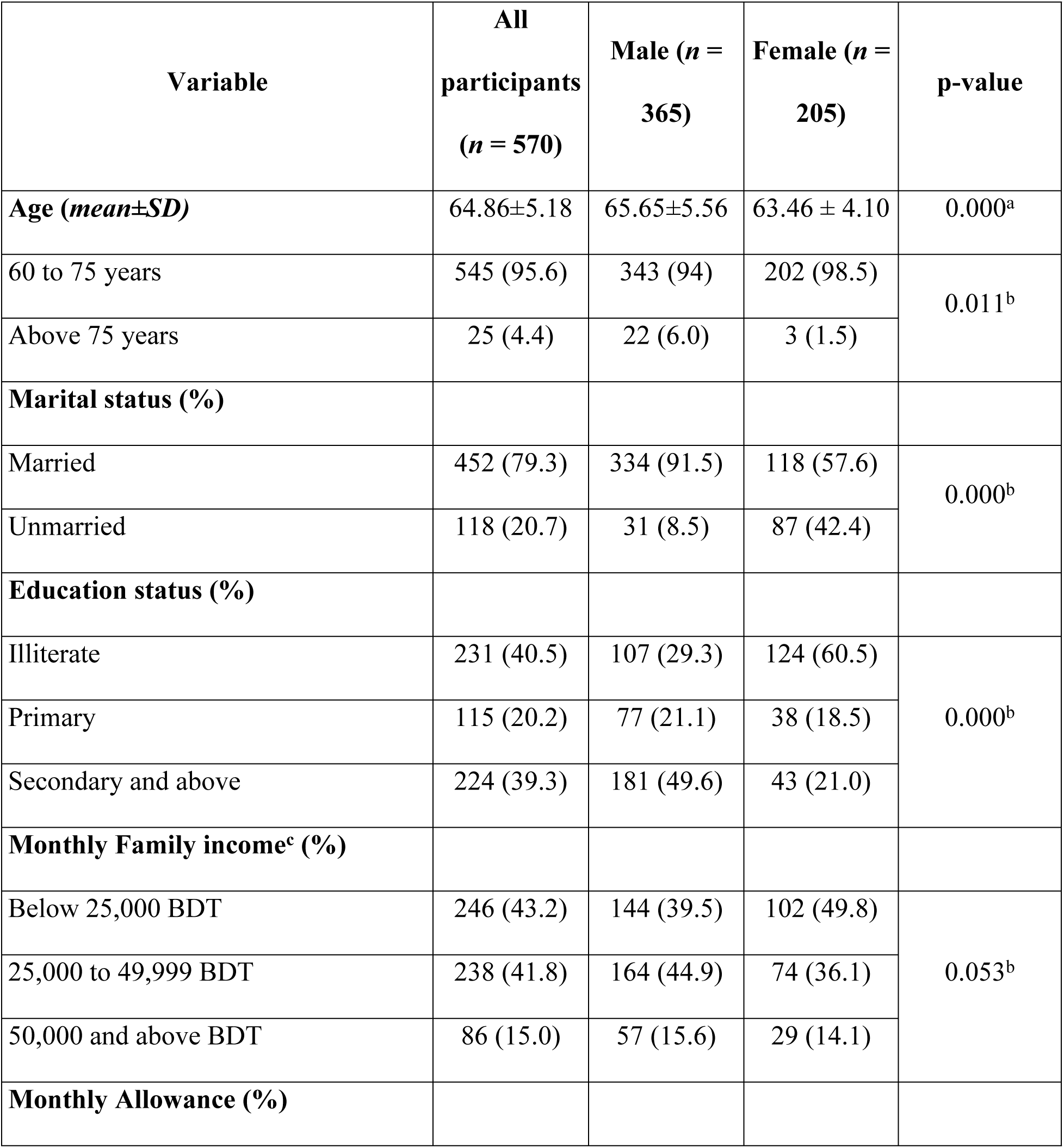
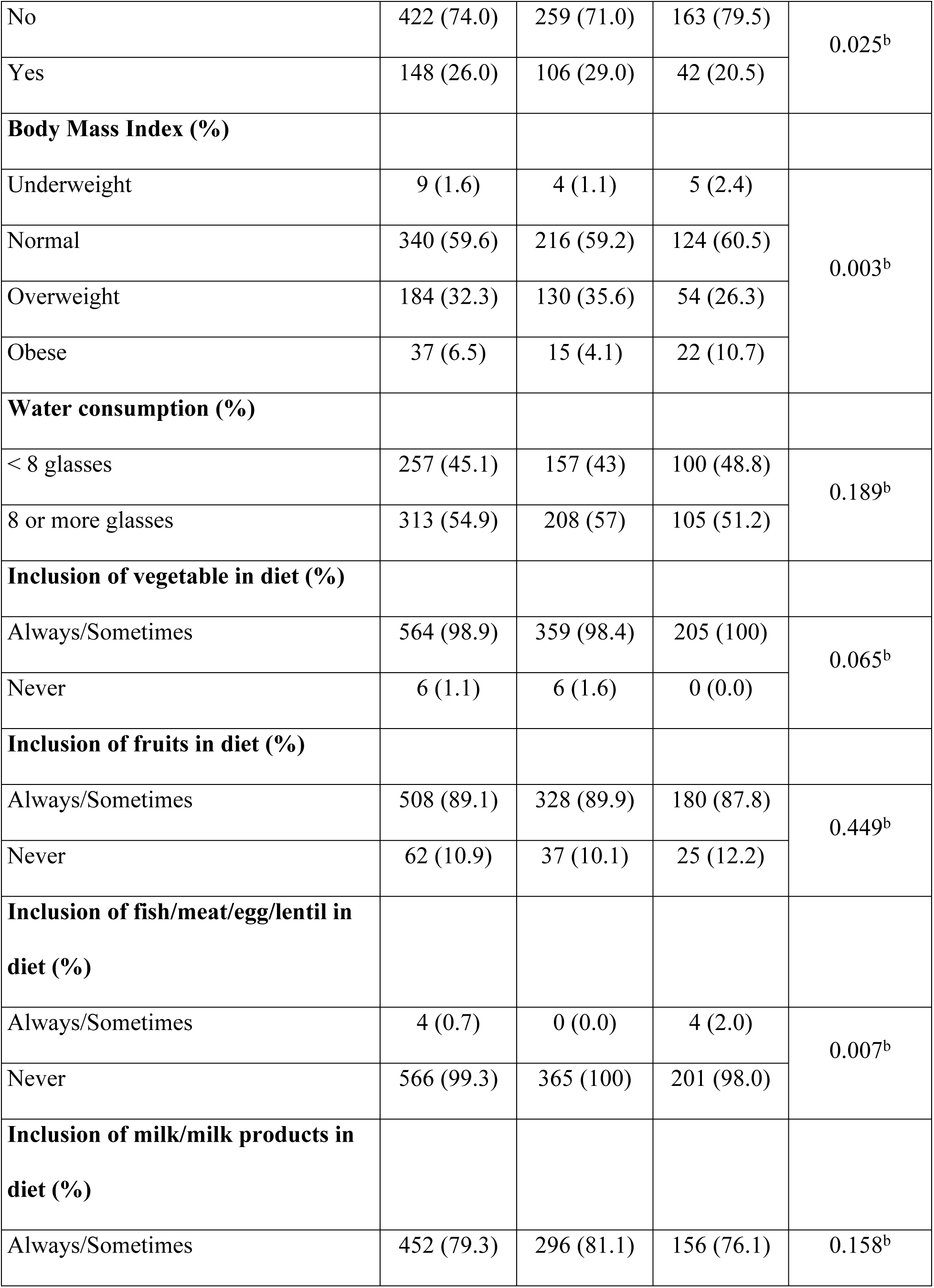
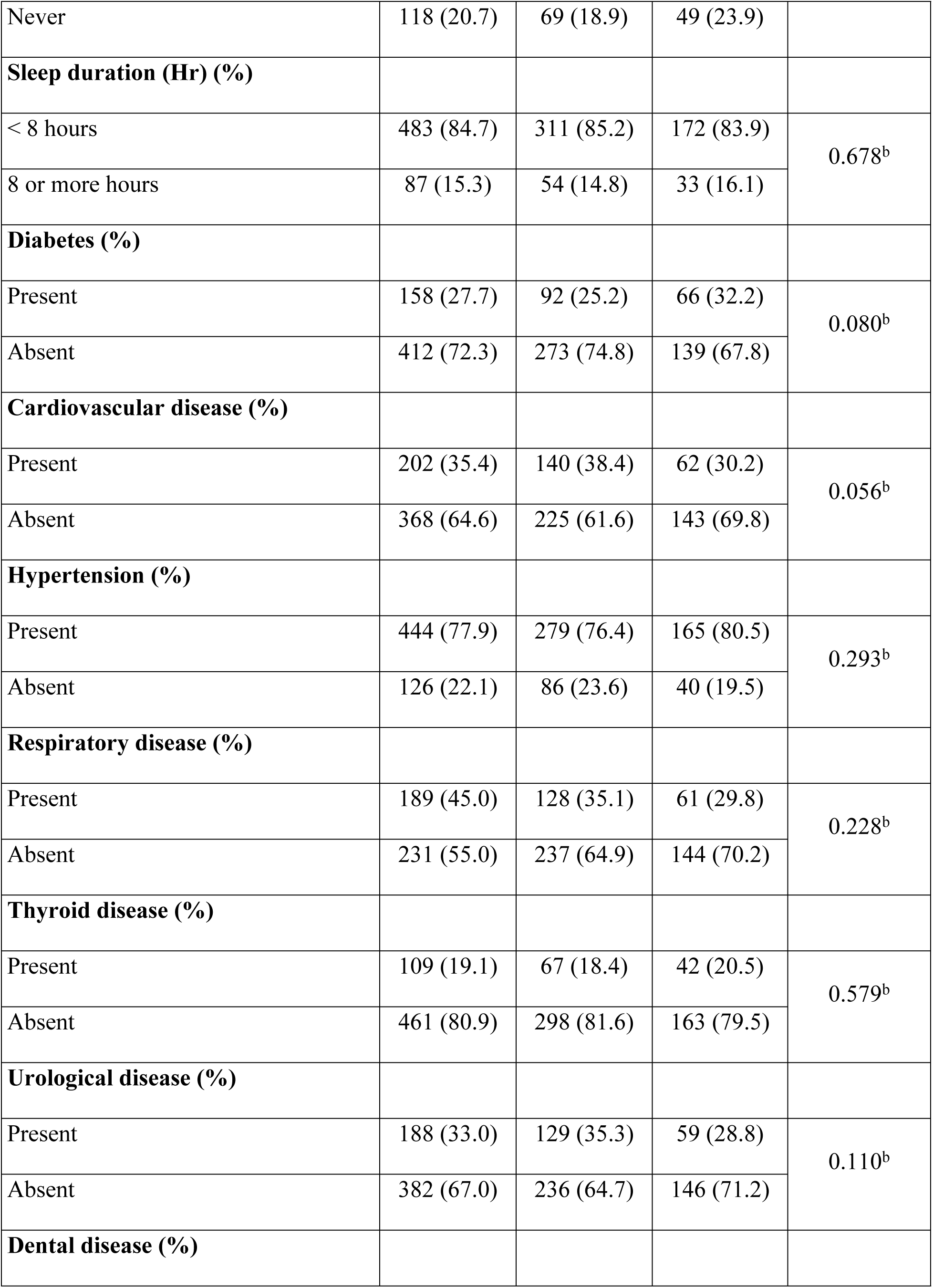
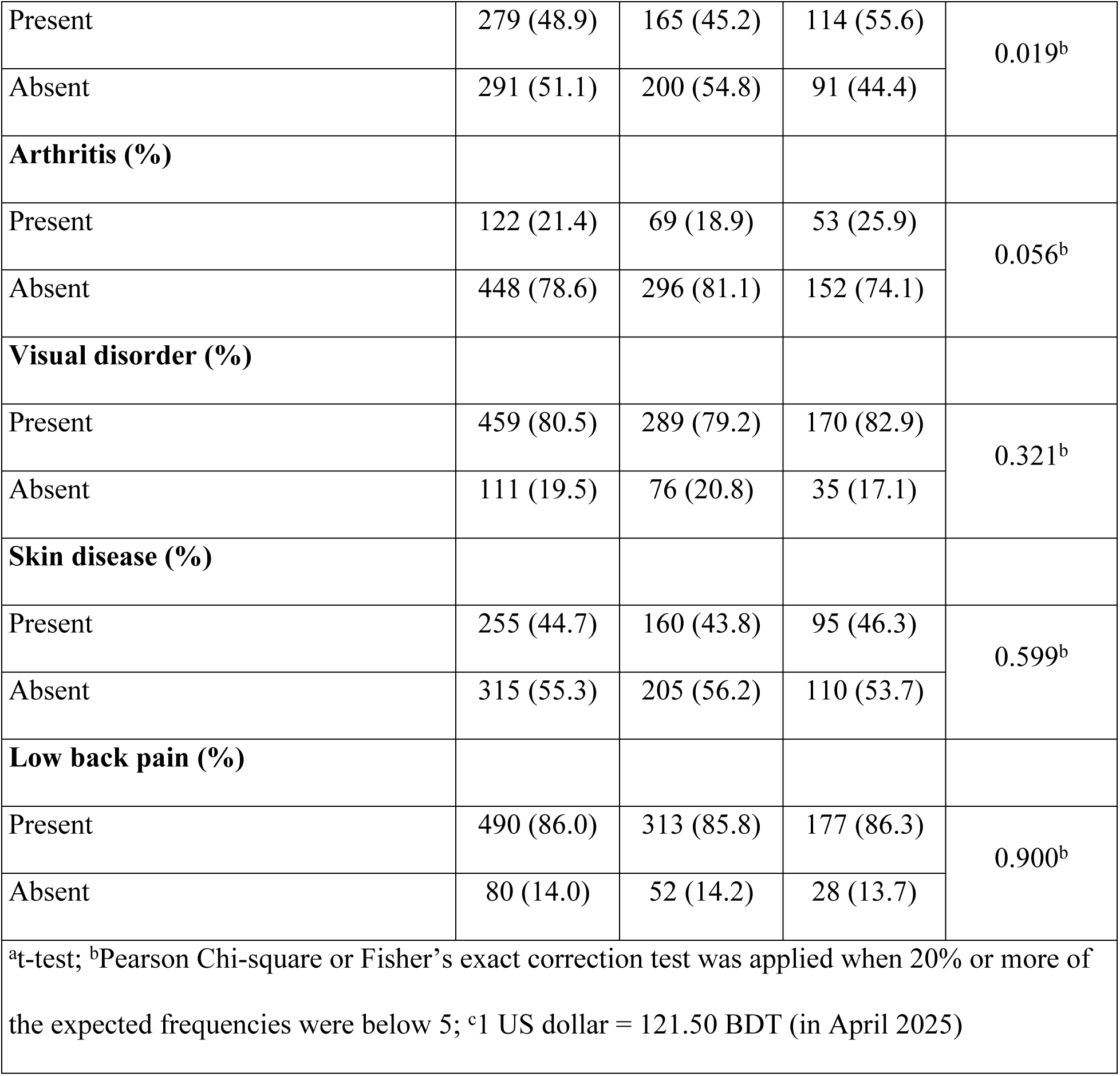
Sociodemographic, physical health, dietary habits & comorbidity status of the elderly participants (*n* = 570)

| Variable | All participants<br>($n = 570$) | Male ($n = 365$) | Female ($n = 205$) | p-value |
| --- | --- | --- | --- | --- |
| **Age (*mean*±*SD*)** | 64.86±5.18 | 65.65±5.56 | 63.46 ± 4.10 | 0.000<sup>a</sup> |
| 60 to 75 years | 545 (95.6) | 343 (94) | 202 (98.5) | 0.011<sup>b</sup> |
| Above 75 years | 25 (4.4) | 22 (6.0) | 3 (1.5) |  |
| **Marital status (%)** |  |  |  |  |
| Married | 452 (79.3) | 334 (91.5) | 118 (57.6) | 0.000<sup>b</sup> |
| Unmarried | 118 (20.7) | 31 (8.5) | 87 (42.4) |  |
| **Education status (%)** |  |  |  |  |
| Illiterate | 231 (40.5) | 107 (29.3) | 124 (60.5) | 0.000<sup>b</sup> |
| Primary | 115 (20.2) | 77 (21.1) | 38 (18.5) |  |
| Secondary and above | 224 (39.3) | 181 (49.6) | 43 (21.0) |  |
| **Monthly Family income<sup>c</sup> (%)** |  |  |  |  |
| Below 25,000 BDT | 246 (43.2) | 144 (39.5) | 102 (49.8) | 0.053<sup>b</sup> |
| 25,000 to 49,999 BDT | 238 (41.8) | 164 (44.9) | 74 (36.1) |  |
| 50,000 and above BDT | 86 (15.0) | 57 (15.6) | 29 (14.1) |  |
| **Monthly Allowance (%)** |  |  |  |  |
| No | 422 (74.0) | 259 (71.0) | 163 (79.5) | 0.025<sup>b</sup> |
| Yes | 148 (26.0) | 106 (29.0) | 42 (20.5) |  |
| **Body Mass Index (%)** |  |  |  |  |
| Underweight | 9 (1.6) | 4 (1.1) | 5 (2.4) | 0.003<sup>b</sup> |
| Normal | 340 (59.6) | 216 (59.2) | 124 (60.5) |  |
| Overweight | 184 (32.3) | 130 (35.6) | 54 (26.3) |  |
| Obese | 37 (6.5) | 15 (4.1) | 22 (10.7) |  |
| **Water consumption (%)** |  |  |  |  |
| < 8 glasses | 257 (45.1) | 157 (43) | 100 (48.8) | 0.189<sup>b</sup> |
| 8 or more glasses | 313 (54.9) | 208 (57) | 105 (51.2) |  |
| **Inclusion of vegetable in diet (%)** |  |  |  |  |
| Always/Sometimes | 564 (98.9) | 359 (98.4) | 205 (100) | 0.065<sup>b</sup> |
| Never | 6 (1.1) | 6 (1.6) | 0 (0.0) |  |
| **Inclusion of fruits in diet (%)** |  |  |  |  |
| Always/Sometimes | 508 (89.1) | 328 (89.9) | 180 (87.8) | 0.449<sup>b</sup> |
| Never | 62 (10.9) | 37 (10.1) | 25 (12.2) |  |
| **Inclusion of fish/meat/egg/lentil in diet (%)** |  |  |  |  |
| Always/Sometimes | 4 (0.7) | 0 (0.0) | 4 (2.0) | 0.007<sup>b</sup> |
| Never | 566 (99.3) | 365 (100) | 201 (98.0) |  |
| **Inclusion of milk/milk products in diet (%)** |  |  |  |  |
| Always/Sometimes | 452 (79.3) | 296 (81.1) | 156 (76.1) | 0.158<sup>b</sup> |
| Never | 118 (20.7) | 69 (18.9) | 49 (23.9) |  |
| **Sleep duration (Hr) (%)** |  |  |  |  |
| < 8 hours | 483 (84.7) | 311 (85.2) | 172 (83.9) | 0.678<sup>b</sup> |
| 8 or more hours | 87 (15.3) | 54 (14.8) | 33 (16.1) |  |
| **Diabetes (%)** |  |  |  |  |
| Present | 158 (27.7) | 92 (25.2) | 66 (32.2) | 0.080<sup>b</sup> |
| Absent | 412 (72.3) | 273 (74.8) | 139 (67.8) |  |
| **Cardiovascular disease (%)** |  |  |  |  |
| Present | 202 (35.4) | 140 (38.4) | 62 (30.2) | 0.056<sup>b</sup> |
| Absent | 368 (64.6) | 225 (61.6) | 143 (69.8) |  |
| **Hypertension (%)** |  |  |  |  |
| Present | 444 (77.9) | 279 (76.4) | 165 (80.5) | 0.293<sup>b</sup> |
| Absent | 126 (22.1) | 86 (23.6) | 40 (19.5) |  |
| **Respiratory disease (%)** |  |  |  |  |
| Present | 189 (45.0) | 128 (35.1) | 61 (29.8) | 0.228<sup>b</sup> |
| Absent | 231 (55.0) | 237 (64.9) | 144 (70.2) |  |
| **Thyroid disease (%)** |  |  |  |  |
| Present | 109 (19.1) | 67 (18.4) | 42 (20.5) | 0.579<sup>b</sup> |
| Absent | 461 (80.9) | 298 (81.6) | 163 (79.5) |  |
| **Urological disease (%)** |  |  |  |  |
| Present | 188 (33.0) | 129 (35.3) | 59 (28.8) | 0.110<sup>b</sup> |
| Absent | 382 (67.0) | 236 (64.7) | 146 (71.2) |  |
| **Dental disease (%)** |  |  |  |  |
| Present | 279 (48.9) | 165 (45.2) | 114 (55.6) | 0.019<sup>b</sup> |
| Absent | 291 (51.1) | 200 (54.8) | 91 (44.4) |  |
| **Arthritis (%)** |  |  |  |  |
| Present | 122 (21.4) | 69 (18.9) | 53 (25.9) | 0.056<sup>b</sup> |
| Absent | 448 (78.6) | 296 (81.1) | 152 (74.1) |  |
| **Visual disorder (%)** |  |  |  |  |
| Present | 459 (80.5) | 289 (79.2) | 170 (82.9) | 0.321<sup>b</sup> |
| Absent | 111 (19.5) | 76 (20.8) | 35 (17.1) |  |
| **Skin disease (%)** |  |  |  |  |
| Present | 255 (44.7) | 160 (43.8) | 95 (46.3) | 0.599<sup>b</sup> |
| Absent | 315 (55.3) | 205 (56.2) | 110 (53.7) |  |
| **Low back pain (%)** |  |  |  |  |
| Present | 490 (86.0) | 313 (85.8) | 177 (86.3) | 0.900<sup>b</sup> |
| Absent | 80 (14.0) | 52 (14.2) | 28 (13.7) |  |
| at-test; bPearson Chi-square or Fisher’s exact correction test was applied when 20% or more of the expected frequencies were below 5; c1 US dollar = 121.50 BDT (in April 2025) |  |  |  |  |

The distribution of Older People Quality of Life (OPQOL) scores among the participants (Figure 1) reveals that most respondents reported a "Bad" quality of life (43.2%), followed by those with an "Optimum" QoL (27.9%). A significant proportion (21.9%) reported their quality of life as "Very Bad" while only a small percentage (7%) reported a "Good" QoL.

**Figure 1.**
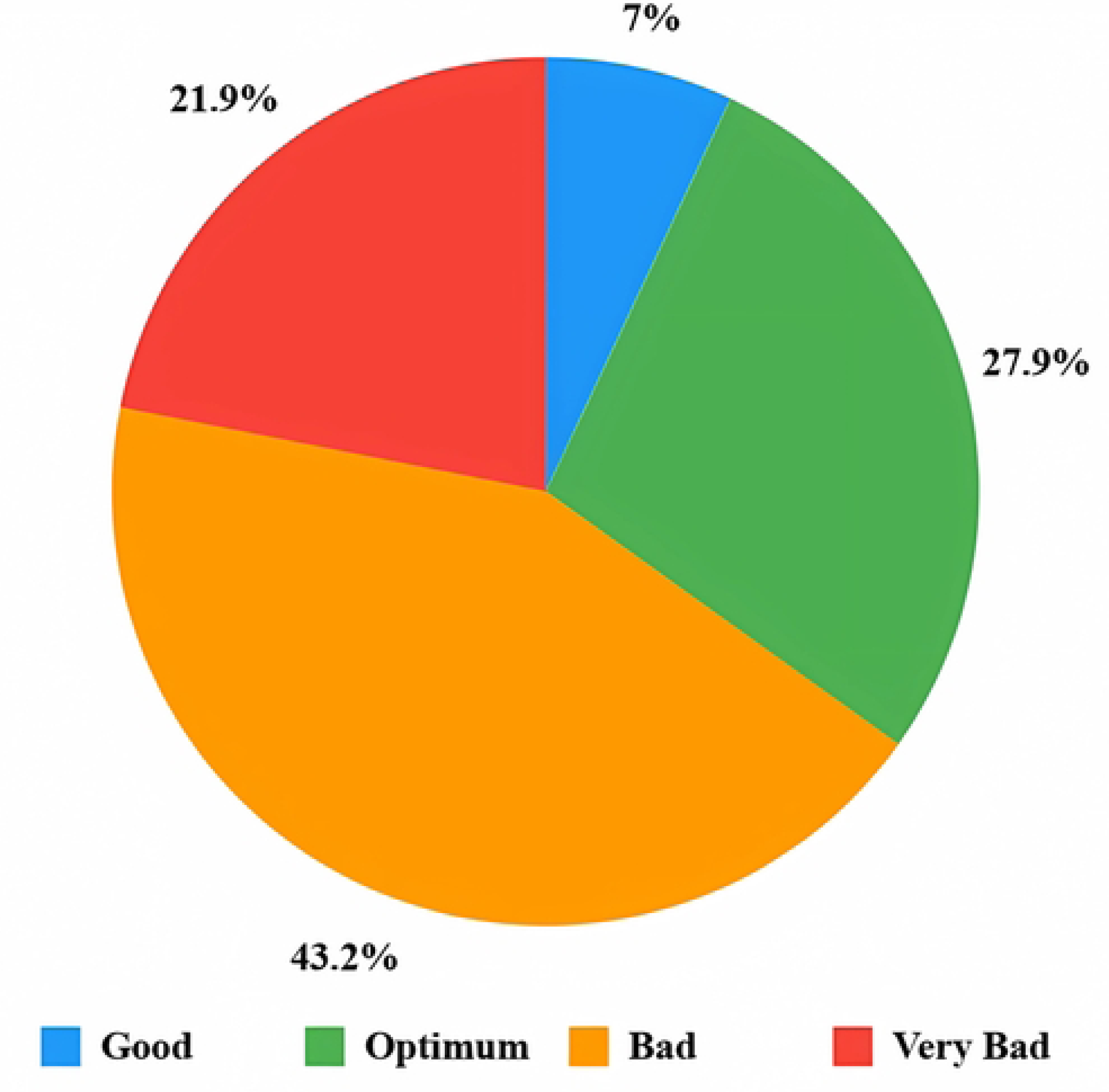
Distribution of OPQOL score among the participants (*n* = 570)

Table 2 further breaks down these findings, showing that the mean OPQOL score for males (116.11 ± 16.36) was significantly higher than that for females (109.33 ± 15.53, p = 0.000). A notably higher percentage of females (30.7%) was seen having "Very Bad" compared to males (17%, p = 0.000). Additionally, while the percentage of males and females reporting a "Bad" QoL was similar (42.7% for males and 43.9% for females, p = 0.000), more males (31.2%) found out to have "Optimum" QoL compared to females (22%, p = 0.000). Furthermore, a higher percentage of males (9%) had a "Good" QoL compared to females (3.4%, p = 0.000), indicating a significant gender disparity in overall quality of life. There were no participants from the category "good as can be".

**Table 2.** Distribution of Quality-of-Life score among the elderly participants (*n* = 570)

| Outcome | All participants<br>(*n* = 570) | Male (*n* = 365) | Female<br>(*n* = 205) | p-value |
| --- | --- | --- | --- | --- |
| Older People Quality of Life (*mean*±*SD*) | 113.67 ±<br>16.38 | 116.11 ±<br>16.36 | 109.33 ±<br>15.53 | 0.000<sup>a</sup> |
| Very Bad | 125 (21.9) | 62 (17.0) | 63 (30.7) | 0.000<sup>b</sup> |
| Bad | 246 (43.2) | 156 (42.7) | 90 (43.9) |  |
| Optimum | 159 (27.9) | 114 (31.2) | 45 (22.0) |  |
| Good | 40 (7.0) | 33 (9.0) | 7 (3.4) |  |
| <sup>a</sup>t-test; <sup>b</sup>Pearson Chi-square test |  |  |  |  |

The logistic regression analysis identified several key predictors of poor quality of life (QoL) among older adults. The results demonstrate that various sociodemographic and health-related factors are significantly associated with QoL after adjusting potential confounders in this population. The dependent variable was the quality of life (0 = good, 1 = bad). The selected independent variables were: age (0 = 60 to 75 years, 1 = above 75 years), gender (0 = male, 1 = female), marital status (married = 0, unmarried= 1), educational status (0 = illiterate, 1 = primary, 2 = secondary and above), monthly family income (0 = below 25,000 BDT, 1 = 25,000 to 49,999 BDT, 2 = 50,000 BDT and above), monthly allowance (0 = Yes, 1= No), sleep duration (0 = 8 or more hours, 1 = < 8 hours), water consumption (0 = 8 or more glasses, 1 = < 8 glasses), inclusion of vegetable in diet (0 = regularly/sometimes, 1 = never), inclusion of meat/fish/egg/lentil in diet (0 = regularly/sometimes, 1 = never), inclusion of fruits in diet (0 = regularly/sometimes, 1 = never), inclusion of milk/milk products in diet (0 = regularly/sometimes, 1 = never), diabetes (0 = present, 1 = absent), cardiovascular disease (0 = present, 1 = absent), hypertension (0 = present, 1 = absent), thyroid disease (0 = present, 1 = absent), skin disease (0 = present, 1 = absent), respiratory disease (0 = present, 1 = absent), dental disease (0 = present, 1 = absent), urological disease (0 = present, 1 = absent), visual disorder (0 = present, 1 = absent), arthritis (0 = present, 1 = absent), low back pain (0 = present, 1 = absent).

**Fig 2:**
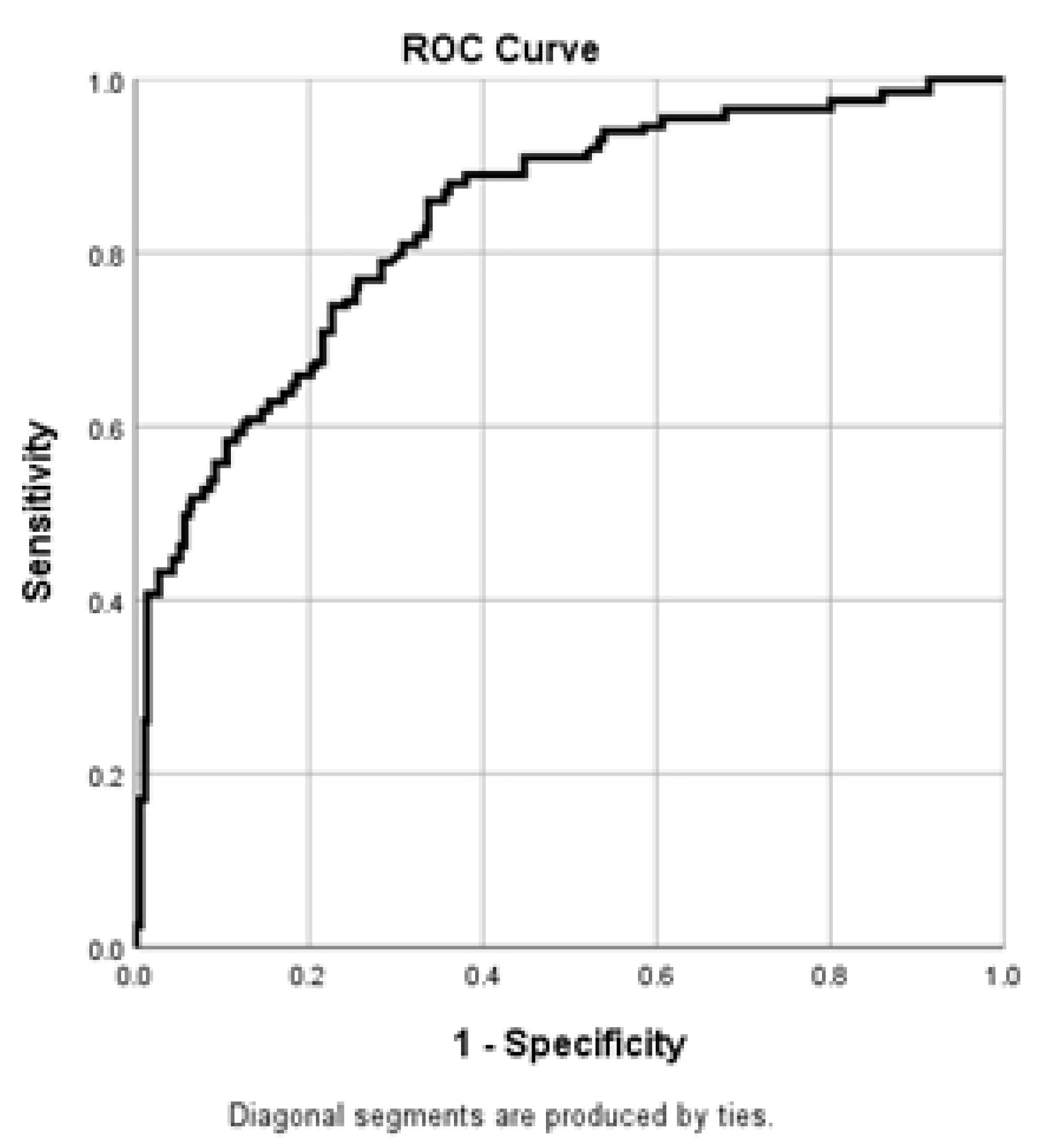
Illustration of ROC curve for the regression model performance.

The model’s overall accuracy in correctly classifying cases was 65.1%. This improved to 77.7% after adding independent variables, indicating an enhanced ability of the model to predict the dependent variable. No multicollinearity was observed. The area under the receiver operating characteristics (ROC) curve was 0.83, with 95% CI ranging from 0.80 to 0.87 and a p-value of 0.000.

In the table 3 it was found that age proved to be an influential determinant, with individuals older than 75 years being 5.14 times more likely to experience poor QoL than individuals between the ages of 60 to 75 years (AOR = 5.14, 95% CI: 1.33-19.86, p = 0.017). Education level was also found to be highly correlated with QoL. Illiterate individuals were found to be 2.50 times more likely to experience a lower QoL than individuals with at least secondary education (AOR = 2.50, 95% CI: 1.43-4.35, p = 0.001). Individuals with an income of less than 25,000 BDT were 10.36 times more likely to report poor QoL than individuals with incomes of 50,000 BDT or higher (AOR = 10.36, 95% CI: 5.10-21.04, p = 0.000). Individuals with incomes between 25,001-49,999 BDT were 7.83 times more likely to report poor QoL (AOR = 7.83, 95% CI: 3.97-15.46, p = 0.000). Individuals not receiving any monthly allowance were 3.14 times more likely to indicate poorer QoL than individuals who received the allowances (AOR = 3.14, 95% CI: 1.87-5.28, p = 0.000). Sleeping less than 8 hours was linked with poorer quality of life (OR = 1.94, 95% CI: 1.06-3.55, p = 0.030). Interestingly, consumption of less than 8 glasses of water per day also turned out to be an independent predictor, with those who consumed less water having more likelihood to report poorer quality of life (OR = 2.18, 95% CI: 1.38-3.43, p = 0.001). Lack of including fruits in diet regularly or often was significantly associated with poorer QoL (OR = 8.01, 95% CI: 2.33-27.49, p = 0.001).

**Table 3.**
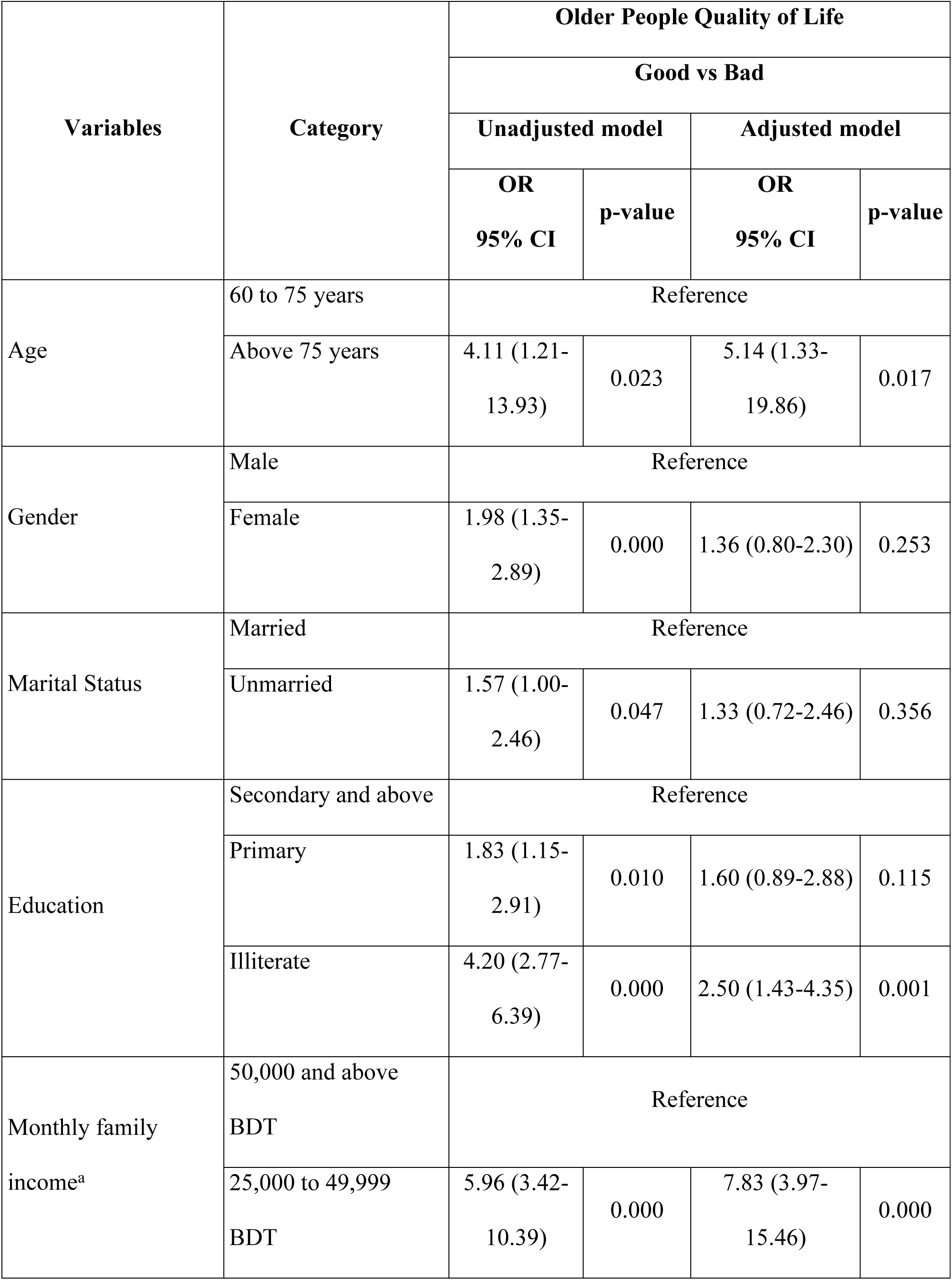
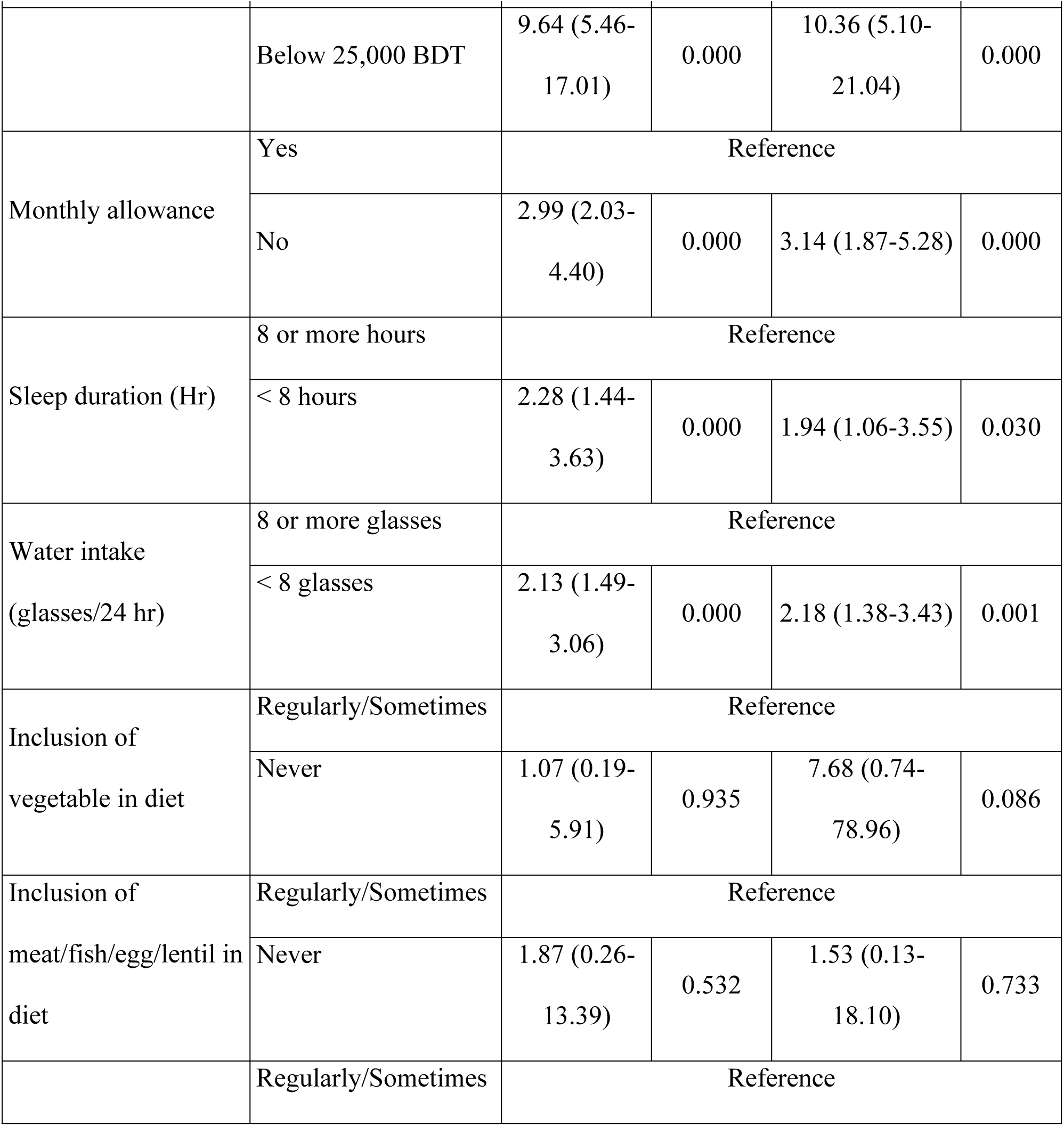
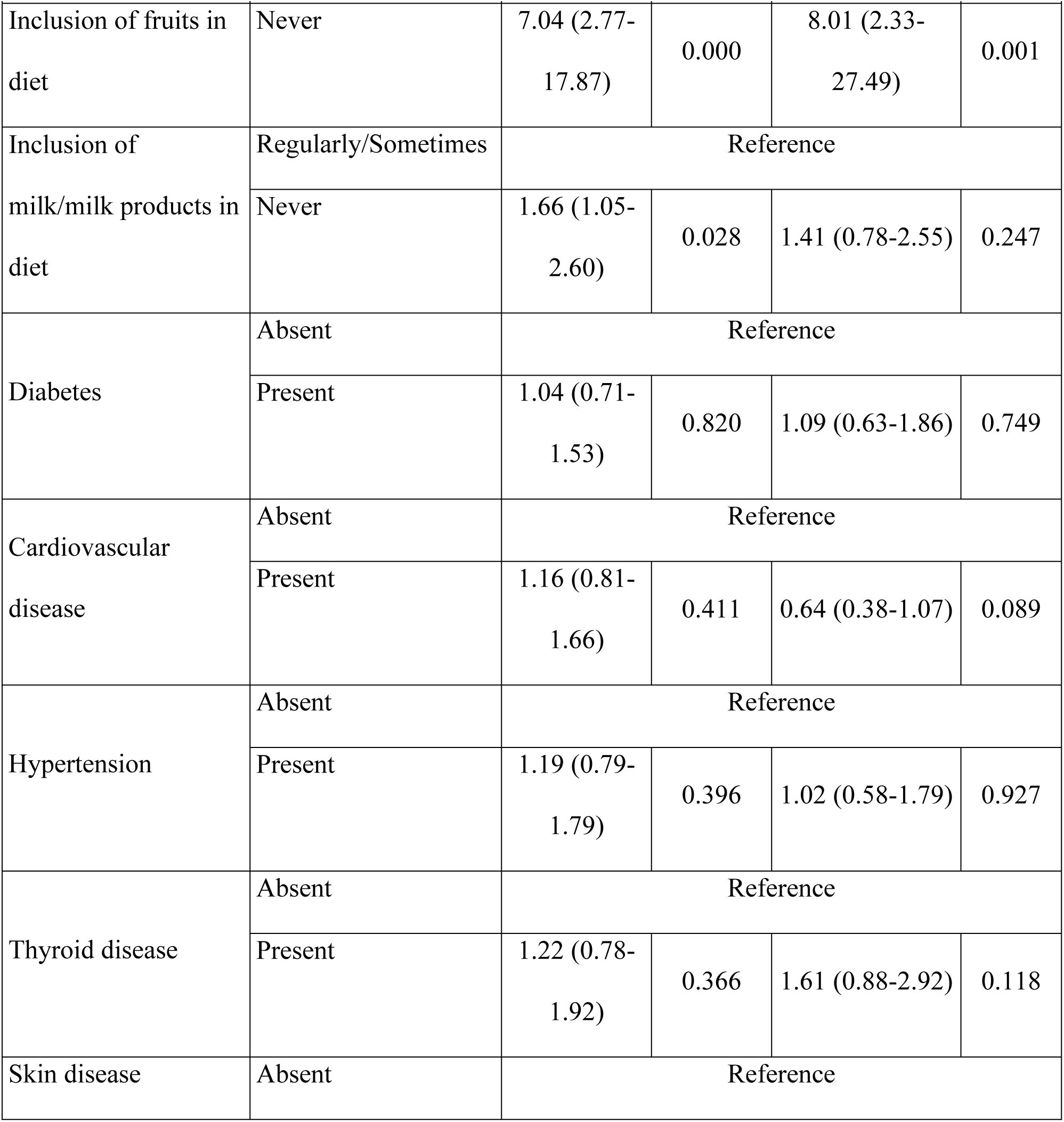
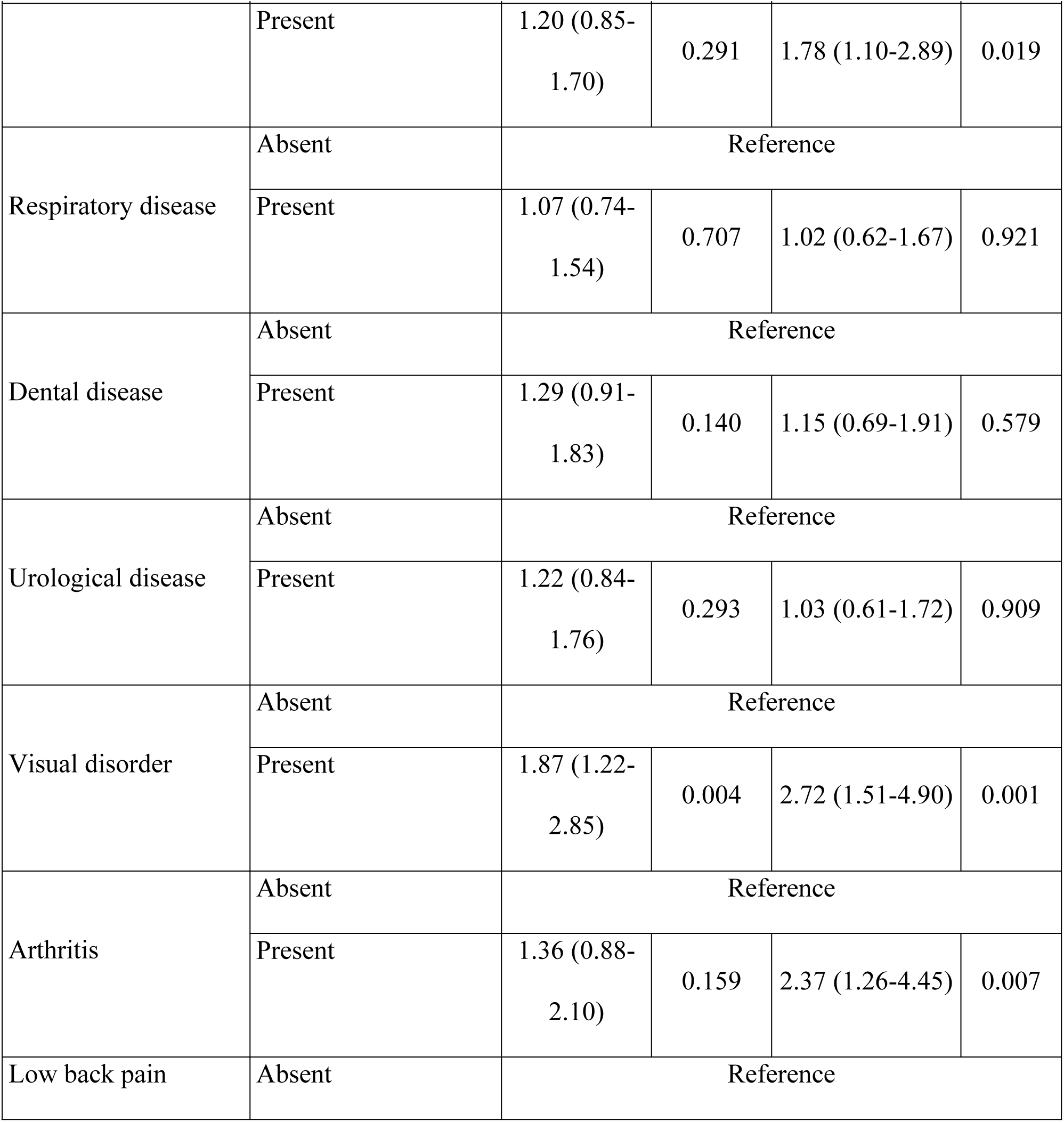
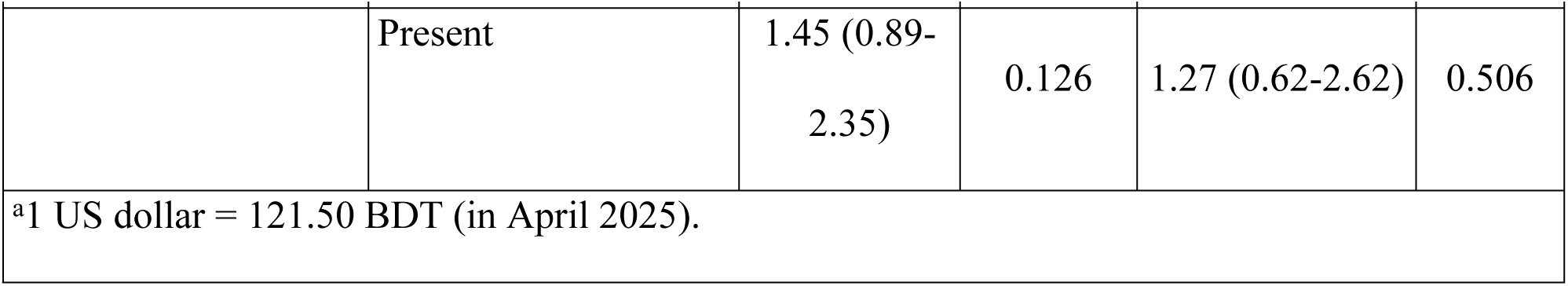
Logistic regression analysis of sociodemographic, physical status, dietary habits & comorbidity status with OPQOL (Multinomial Logistic regression)

| Variables | Category | Older People Quality of Life |  |  |  |
| --- | --- | --- | --- | --- | --- |
|  |  | Good vs Bad |  |  |  |
|  |  | Unadjusted model |  | Adjusted model |  |
|  |  | OR<br>95% CI | p-value | OR<br>95% CI | p-value |
| Age | 60 to 75 years | Reference |  |  |  |
|  | Above 75 years | 4.11 (1.21-13.93) | 0.023 | 5.14 (1.33-19.86) | 0.017 |
| Gender | Male | Reference |  |  |  |
|  | Female | 1.98 (1.35-2.89) | 0.000 | 1.36 (0.80-2.30) | 0.253 |
| Marital Status | Married | Reference |  |  |  |
|  | Unmarried | 1.57 (1.00-2.46) | 0.047 | 1.33 (0.72-2.46) | 0.356 |
| Education | Secondary and above | Reference |  |  |  |
|  | Primary | 1.83 (1.15-2.91) | 0.010 | 1.60 (0.89-2.88) | 0.115 |
|  | Illiterate | 4.20 (2.77-6.39) | 0.000 | 2.50 (1.43-4.35) | 0.001 |
| Monthly family income<sup>a</sup> | 50,000 and above BDT | Reference |  |  |  |
|  | 25,000 to 49,999 BDT | 5.96 (3.42-10.39) | 0.000 | 7.83 (3.97-15.46) | 0.000 |
|  |  | Good vs Bad |  |  |  |
|  |  | Unadjusted model |  | Adjusted model |  |
|  |  | OR<br>95% CI | p-value | OR<br>95% CI | p-value |
|  | Below 25,000 BDT | 9.64 (5.46-17.01) | 0.000 | 10.36 (5.10-21.04) | 0.000 |
| Monthly allowance | Yes | Reference |  |  |  |
|  | No | 2.99 (2.03-4.40) | 0.000 | 3.14 (1.87-5.28) | 0.000 |
| Sleep duration (Hr) | 8 or more hours | Reference |  |  |  |
|  | < 8 hours | 2.28 (1.44-3.63) | 0.000 | 1.94 (1.06-3.55) | 0.030 |
| Water intake<br>(glasses/24 hr) | 8 or more glasses | Reference |  |  |  |
|  | < 8 glasses | 2.13 (1.49-3.06) | 0.000 | 2.18 (1.38-3.43) | 0.001 |
| Inclusion of<br>vegetable in diet | Regularly/Sometimes | Reference |  |  |  |
|  | Never | 1.07 (0.19-5.91) | 0.935 | 7.68 (0.74-78.96) | 0.086 |
| Inclusion of<br>meat/fish/egg/lentil in<br>diet | Regularly/Sometimes | Reference |  |  |  |
|  | Never | 1.87 (0.26-13.39) | 0.532 | 1.53 (0.13-18.10) | 0.733 |
|  | Regularly/Sometimes | Reference |  |  |  |
|  |  | Good vs Bad |  |  |  |
|  |  | Unadjusted model |  | Adjusted model |  |
|  |  | OR<br>95% CI | p-value | OR<br>95% CI | p-value |
| Inclusion of fruits in diet | Never | 7.04 (2.77-17.87) | 0.000 | 8.01 (2.33-27.49) | 0.001 |
| Inclusion of milk/milk products in diet | Regularly/Sometimes | Reference |  |  |  |
|  | Never | 1.66 (1.05-2.60) | 0.028 | 1.41 (0.78-2.55) | 0.247 |
| Diabetes | Absent | Reference |  |  |  |
|  | Present | 1.04 (0.71-1.53) | 0.820 | 1.09 (0.63-1.86) | 0.749 |
| Cardiovascular disease | Absent | Reference |  |  |  |
|  | Present | 1.16 (0.81-1.66) | 0.411 | 0.64 (0.38-1.07) | 0.089 |
| Hypertension | Absent | Reference |  |  |  |
|  | Present | 1.19 (0.79-1.79) | 0.396 | 1.02 (0.58-1.79) | 0.927 |
| Thyroid disease | Absent | Reference |  |  |  |
|  | Present | 1.22 (0.78-1.92) | 0.366 | 1.61 (0.88-2.92) | 0.118 |
| Skin disease | Absent | Reference |  |  |  |
|  |  | Good vs Bad |  |  |  |
|  |  | Unadjusted model |  | Adjusted model |  |
|  |  | OR<br>95% CI | p-value | OR<br>95% CI | p-value |
|  | Present | 1.20 (0.85-1.70) | 0.291 | 1.78 (1.10-2.89) | 0.019 |
| Respiratory disease | Absent | Reference |  |  |  |
|  | Present | 1.07 (0.74-1.54) | 0.707 | 1.02 (0.62-1.67) | 0.921 |
| Dental disease | Absent | Reference |  |  |  |
|  | Present | 1.29 (0.91-1.83) | 0.140 | 1.15 (0.69-1.91) | 0.579 |
| Urological disease | Absent | Reference |  |  |  |
|  | Present | 1.22 (0.84-1.76) | 0.293 | 1.03 (0.61-1.72) | 0.909 |
| Visual disorder | Absent | Reference |  |  |  |
|  | Present | 1.87 (1.22-2.85) | 0.004 | 2.72 (1.51-4.90) | 0.001 |
| Arthritis | Absent | Reference |  |  |  |
|  | Present | 1.36 (0.88-2.10) | 0.159 | 2.37 (1.26-4.45) | 0.007 |
| Low back pain | Absent | Reference |  |  |  |
|  |  | Good vs Bad |  |  |  |
|  |  | Unadjusted model |  | Adjusted model |  |
|  |  | OR<br>95% CI | p-value | OR<br>95% CI | p-value |
|  | Present | 1.45 (0.89-2.35) | 0.126 | 1.27 (0.62-2.62) | 0.506 |
| <sup>a</sup>1 US dollar = 121.50 BDT (in April 2025). |  |  |  |  |  |

In regard to health condition, individuals with skin disease had a 1.78 times greater likelihood of reporting poorer QoL compared to the absence of skin disease (OR = 1.78, 95% CI: 1.10-2.89, p = 0.019). Visual disorders (OR = 2.72, 95% CI: 1.51-4.90, p = 0.001) also had an effect on QoL. In addition, individuals with arthritis had 2.37 times greater likelihood of reporting poorer QoL than individuals free of arthritis (OR = 2.37, 95% CI: 1.26-4.45, p = 0.007).

No multicollinearity existed between the independent variables. It has been observed that poor QoL among institutionalized elderly Bangladeshis is highly determined by sociodemographic variables like age, education, income, monthly allowance along with factors related to physical health and dietary habits like sleep, water, fruit intake, comorbidity like skin disease, visual disorders, and arthritis.

## Discussion

The objective of this research was to determine the determinants that affect the quality of life (QoL) of elderly individuals at old homes in Bangladesh, based specifically on the demographic, healthcare-related, dietary and comorbidity determinants. This research helps to make known how these determinants interact to affect the well-being of elderly populations residing in institutions, while contributing to the formulation of appropriate policies and interventions.

In this research age, was the most predictive determinant of QoL. Participants older than the age of 75 years were found to be 5.14 times more likely to experience an unsatisfactory QoL than those between the ages of 60–75. This is consistent with the overall pattern, seen in the study of aging that suggests deterioration in physical and mental well-being with aging, with heightened susceptibility to diseases like cardiovascular diseases, diabetes, and arthritis [16,17]. Our results emphasized the need to focus the specific requirements of this elderly group in order to improve their quality of life.

The research further revealed that the females were more vulnerable to illiteracy, where 60.5% of females lacked the ability to read or write compared to 29.3% of the males. This education gap is an important determiner of QoL since lower levels of education contribute to lower levels of health literacy, limiting access to healthcare services, which further impacts the management of diseases and adopting preventive behaviors [18,19]. This is further supported by our findings, which indicated that the illiterate respondents were 2.50 times more likely to describe having poorer QoL than respondents with secondary or higher levels of education.

Economic considerations also significantly contributed to QoL, where individuals who earned less than 25,000 BDT per month were found to be 10.36 times more likely to experience lower QoL than those with incomes of 50,000 BDT or higher. These results are supported by studies that revealed that financial insecurity among the elderly enhances reliance on the family and increases susceptibility to abuse and neglect, ultimately reducing the quality of life [20]. A large number of the Bangladesh respondents received no monthly allowance, more so among females 79.5% compared to 71% of the males, which would likely be responsible for the lower scores of QoL among them.

Sufficient hydration and good sleep were also linked to improved QoL because dehydration and lack of sleep can worsen the progression of underlying health, lead to cognitive decline, mobility limitation, and social isolation [21,22]. These factors are also found to be significantly responsible in this study. Elderly individuals were 1.94 times and 2.18 prone to bad QoL due to lack of 8 hours sleep and drinking water less than 8 glass everyday respectively.

Regarding dietary habit, diet was also seen to be an important modifiable determinant that affected QoL. Inclusion in the diet of fruits, vegetables, and high-protein foods was found to positively impact QoL by maintaining muscle mass, ensuring cognitive abilities, and reducing chronic disease risk [23,24]. Therefor the study here reported significant associations with fruit consumption where lack of inclusion of fruits in diet can increase the chance of poor QoL by 8 times. Similar studies have indicated that increased consumption of fruits and vegetables can delay cognitive decline and even prevent dementia [25,26]. Additionally, consumption of dairy products have been associated with enhanced bone health and cognitive performance, which play a crucial role in sustaining independence in older years [27,28].

Our research concurs with existing research that has documented the adverse effect on older adults’ quality of life (QoL). Studies have proven that the incidence of itching rises with age, and occurs in 20.8% of people between the ages of 60-69 years, 22.9% between the ages of 70-79 years, and 26% aged 75 years and more [29]. Studies have also determined that a significant number of institutionalized elderly subjects reported itching for less than 6 hours a day, but even this short period of time adversely affected the quality of life [29]. This study has also scrutinized the association of skin disease with QoL and found to that skin diseases can impact 1.78-time poor quality of life among elderly individuals.

Brown and Barrett, as well as Wang, found the association between visual impairment and reduced life satisfaction, in keeping similar with our finding that compromised vision negatively affects QoL [30,31]. Furthermore, Marback found that binocular loss of sight has an even deeper impact on QoL. Consistent with other research, our results suggest that visual impairment has a considerable impact on the quality of life (QoL) in the older adult [32]. In our study, visual disorders were highly correlated to reduced QoL, in keeping with other studies that noted an increased risk for adverse health outcomes. This finding shows that the severity of vision impairment matters. It should be considered when creating interventions. This will help improve the quality of life for older adults with visual disorders. It ensures better support for those with severe vision loss.

In our adjusted analysis, poorer QoL was associated with arthritis, in agreement with Dominick’s finding that older adults with arthritis consistently report poorer general health and physical health compared with their non-arthritis counterparts [33]. In addition, our analysis implies that experiencing more chronic diseases further accelerates the loss in QoL. This is in support of another study in the USA, where participants with both arthritis and other chronic illnesses like heart disease, diabetes, or hypertension have an even more aggravated decline in QoL [34]. These results emphasize the synergistic impact of co-morbidities on health outcomes as well as stress the importance of management strategies that address multiple disease states in older adults in order to enhance their overall QoL.

Overall, this research emphasizes multiple facets influencing older adults’ QoL in terms of sociodemographic, health, life-style and comorbidity variables. The results emphasize the need for gender inequality to be addressed, healthcare accessibility to be enhanced, along with encouraging health-promoting life-styles like physical fitness, healthful nutrition, and sufficient sleep. With an emphasis placed on these determinants, strategies can be formulated to enhance the QoL among older people in care facilities and to make aging more healthful and meaningful.

## Conclusion

The present study determined some determinants of older adults’ quality of life (QoL) in Bangladesh, emphasizing sociodemographic variables (age, education, income, monthly allowance) along with factors related to physical health and dietary habits like sleep, water, fruit intake, comorbidity like skin disease, visual disorders, and arthritis in influencing elder’s quality of life. The findings confirmed that poorer education, chronic health conditions, and poorer income levels were highly related to lower QoL, emphasizing that targeted interventions in these areas, especially in an institutional context, need to be addressed. In future research, longitudinal designs should be employed to allow more accurate assessment of these factors’ long-term influence, with objective health measures included in combination with QoL.

## Data Availability

All relevant data are within the manuscript and its Supporting Information files.

## Acknowledgement

We would like to thank Muhammad Millat Hossain, Associate professor, Bangladesh Health Professions Institute, Bangladesh who helped us through this study.

## Author Contribution

**Conceptualization:** Pradip Kumar Saha, Dr. Mohima Sharmin

**Data curation:** Tasrima Trisha Ratna, Somaiya Islam, Kashfia Rahman

**Formal analysis:** Masum Mridha, Shahriar Hasan

**Investigation:** Tasrima Trisha Ratna, Masum Mridha, Shahriar Hasan

**Methodology:** Dr. Mohima Sharmin, Somaiya Islam

**Project administration:** Pradip Kumar Saha, G M Mainuddin Chisty

**Resources:** Joynal Abedin Imran, Umme Salma, Kashfia Rahman

**Software:** Somaiya Islam, Shahriar Hasan

**Supervision:** Dr. Mohima Sharmin, Pradip Kumar Saha

**Validation:** Pradip Kumar Saha, G M Mainuddin Chisty

**Visualization:** Shahriar Hasan, Pradip Kumar Saha

**Writing - original draft:** Tasrima Trisha Ratna, Somaiya Islam

**Writing - review & editing:** Masum Mridha, Umme Salma

## Supporting information

**S1 Informed consent paper.** The included written consent form administered to the participants.

**S2 Questionnaire.** The included questions comprised the questionnaire administered to the participants.

**S3 Data.** The included data of the participants.

